# Epigenome-wide association study meta-analysis of BMI in African American Adults

**DOI:** 10.1101/2025.01.15.25320607

**Authors:** Kendra Ferrier, Mariaelisa Graff, Iain R. Konigsberg, Maggie Stanislawski, Heather M. Highland, Laura M. Raffield, April P. Carson, Eric Boerwinkle, Jill M. Norris, Chris R. Gignoux, Audrey E. Hendricks, Sridharan Raghavan, Kari E. North, Matthew A. Allison, Mathew J. Budoff, Silva Kasela, François Aguet, Joshua J. Joseph, Charles Kooperberg, Stephen S. Rich, Jerome I. Rotter, Ethan M. Lange, Leslie A. Lange

**Affiliations:** Department of Biomedical Informatics, University of Colorado Anschutz Medical Campus, Aurora, CO, USA; Department of Epidemiology, University of North Carolina, Chapel Hill, NC, United States of America; Department of Genetics, University of North Carolina, Chapel Hill, NC, USA; Department of Medicine, University of Mississippi Medical Center, Jackson, MS, USA; Human Genetics Center, School of Public Health, The University of Texas Health Science Center, Houston, TX, USA; Department of Epidemiology, Colorado School of Public Health, Aurora, CO, USA; Department of Medicine, University of Colorado Anschutz Medical Campus, Aurora, CO, USA; Department of Family Medicine, University of California San Diego, La Jolla, CA, USA; Division of Cardiology, The Lundquist Institute for Biomedical Innovation at Harbor-UCLA Medical Center, Torrance, CA, USA; New York Genome Center, New York, NY, USA; The Broad Institute of MIT and Harvard, Cambridge, MA, USA; Department of Medicine, The Ohio State University Wexner Medical Center, Columbus, OH, USA; Division of Public Health Sciences, Fred Hutchinson Cancer Center, Seattle, WA, USA; Center for Public Health Genomics, Department of Public Health Sciences, University of Virginia, Charlottesville, VA, USA; The Institute for Translational Genomics and Population Sciences, Department of Pediatrics, The Lundquist Institute for Biomedical Innovation at Harbor-UCLA Medical Center, Torrance, CA, USA

## Abstract

Despite considerable advances in identifying risk factors for obesity development, there remains substantial gaps in our knowledge about its etiology. Variation in obesity (defined by BMI) is thought to be due in part to heritable factors; however, obesity-associated genetic variants only account for a small portion of heritability. Epigenetic regulation defined by genetic and/or environmental factors with changes in gene expression, may account for some of this “missing heritability”. Epigenetic studies of obesity have largely been conducted in populations of European ancestry, despite the disproportionate burden of obesity in African Americans (AAs). To address race/ethnic (RE)-differences in obesity, we conducted a BMI epigenome-wide association study (EWAS) meta-analysis using AA participants from the Jackson Heart Study (JHS, n=1604) and the Multi-Ethnic Study of Atherosclerosis (MESA, n=179). Analyses using a linear regression model with methylation as the outcome and continuous BMI as the predictor were stratified by study and sex, then meta-analyzed. There were 208 methylation sites (CpGs) that reached epigenome-wide significance (p< 8.72x10^-8^); 151 of these were novel. Of the novel CpGs, 29 CpGs were available for replication testing in a separate sample of AA and 20 replicated. Differentially methylated region (DMR) analysis resulted in 54 DMRs significantly associated with BMI. Several regions are proximal to, or include, genes previously associated with obesity traits (e.g., *SOCS3*, *ABCG1*, and *TGFB1*) in GWAS. Gene and trait enrichment and pathway analysis showed enrichment for genes in immune system and inflammation related pathways (e.g., the IL-6/JAK/STAT pathway). In conclusion, EWAS of BMI in AAs replicated previously known associations identified in European and multi-ethnic EWAS and identified novel obesity-associated CpGs.

## INTRODUCTION

The global prevalence of obesity has nearly tripled in the last 50 years (World Health Organization, 2020). While a rise in obesity prevalence has been observed across all populations in the United States, some racial and ethnic groups are disproportionately affected by obesity yet are not well represented in medical research(Hales et al., 2020). In 2017-2018, the prevalence of obesity was the highest in non-Hispanic African American (AA) adults (49.6%), as compared to non-Hispanic European American (EA) adults (42.4%), Hispanic American adults (44.8%), and non-Hispanic Asian American adults (17.4%) (Hales et al., 2020). Studies of obesity specifically in AA populations are necessary to begin to address the underrepresentation of this high-risk population in medical research and to help identify if there are race/ethnicity (RE)-specific factors contributing to the development of obesity.

Evidence from twin, family, and adoption studies supports that 40-70% of the variation in obesity is due to heritable genetic factors (Albuquerque et al., 2016; Diels et al., 2020). Contrarily, not everyone with increased genetic risk of obesity becomes obese. Research integrating genetic and methylation data suggests that many genetic variant associations with obesity, in part, be mediated by epigenetic regulation (Diels et al., 2020). However genetic and epigenetic studies of obesity have mainly been conducted in individuals with European genetic ancestry. It has been well established that differences in genetic architecture across RE groups can influence genetic predisposition to disease, and evidence supports that like genetic data, epigenetic data also mirrors genetic ancestry information (Bentley et al., 2017; Rahmani et al., 2017). While there are certainly shared obesity genetic variants across different RE populations, minor allele frequencies and association effect sizes can vary drastically between them (Choquet et al., 2013; Hassanein et al., 2010; Loos & Yeo, 2014). Underscoring that methylation is also influenced by environment, there is evidence that exposure to persistent social and economic adversity, an experience that is shared among many AAs, is associated with accelerated epigenetic aging and consequently greater risk of developing early chronic illnesses (Simons et al., 2021). Thus, epigenetic studies of obesity in AA populations could help clarify the contributions of genetic background and environmental exposures to obesity in a disproportionately burdened RE minority population.

There are also known sex differences in obesity prevalence; for example, it has been observed that non-Hispanic AA females have the highest prevalence of obesity, at 56.9%, as compared to other RE groups (Hales et al., 2020). Results from previous epidemiologic studies of obesity support that there are sex differences in lipid metabolism that are due to genetic and hormonal differences that contribute to an increased susceptibility for females to develop obesity as compared to males (Link & Reue, 2017). Sex-stratified genome-wide association studies (GWAS) of obesity-related traits have repeatedly shown that more genetic risk variant associations are identified, and those that are identified have larger effect sizes, in females as compared to males. There are also likely sex-based societal/environmental differences that contribute to obesity risk via epigenetic regulation, but such factors can’t be easily disentangled when data from males and females are directly combined in EWAS analyses. To date, there are few epigenetic studies of obesity that stratify by sex, which makes it more difficult to identify sex-differentiated methylation signals associated with obesity.

In this study, we performed a meta-analysis of sex-stratified BMI epigenome-wide association study (EWAS) results from AAs using data from participants from the Jackson Heart Study (JHS) and the Multi-Ethnic Study of Atherosclerosis (MESA) to identify epigenetic variants associated with obesity in this population. We also used data from AA participants from the Women’s Health Initiative (WHI) and the Atherosclerosis Risk in Communities Study (ARIC) to assess replication of novel BMI epigenetic associations for the subset of CpGs that were measured across all four studies. Additionally, we performed differentially methylated region (DMR) analysis and evaluated the potential functional impacts of our findings using trait, gene, and pathway enrichment analysis. A summary of the analyses conducted in this study is shown in Figure 1.

**Figure 1:**
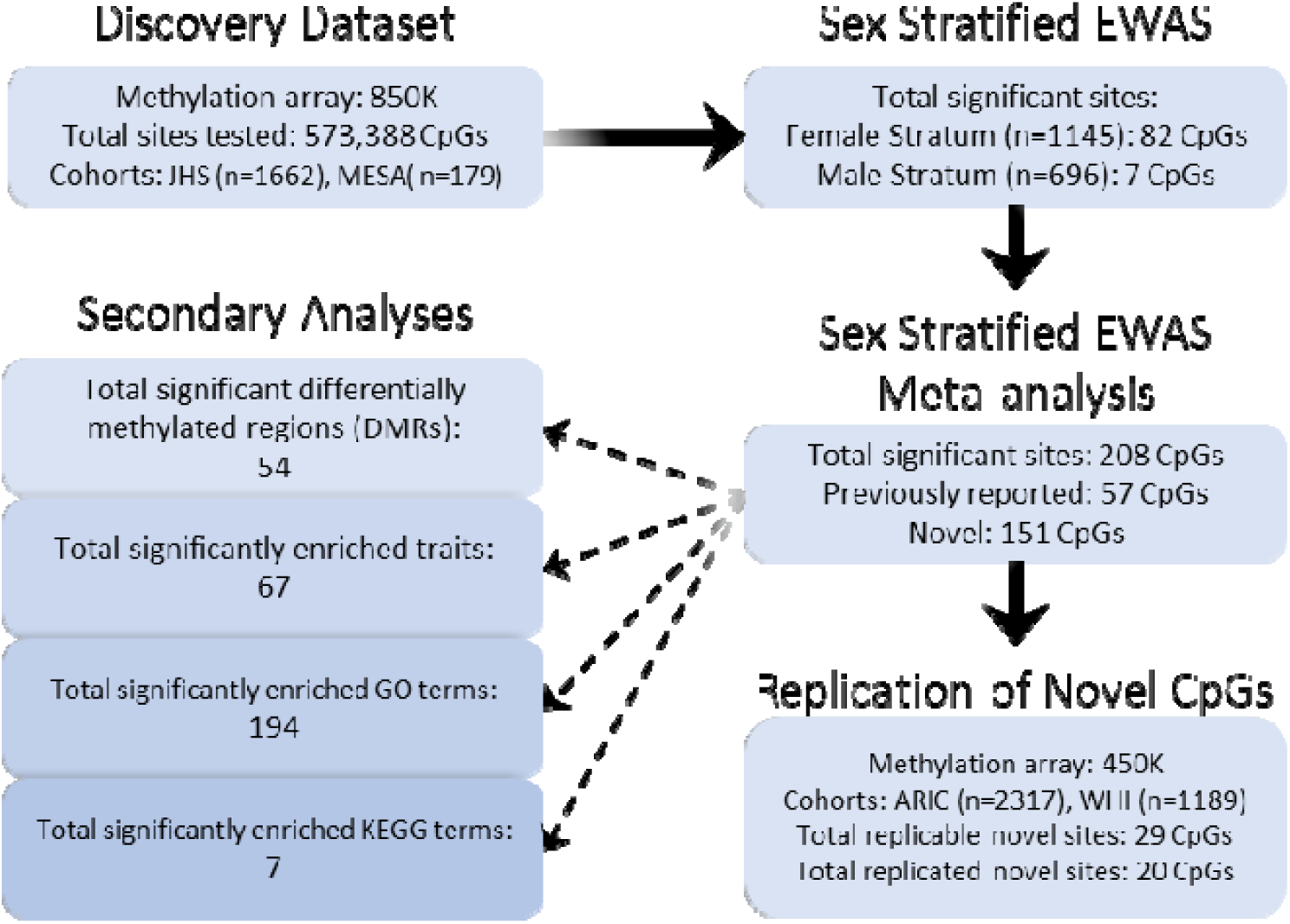
Summary of analyses and results.

## 2. METHODS

### Cohorts

A summary of demographic information for each cohort can be found in Table 1.

**Table 1:** Demographic information of participants. Data is shown by mean (sd) or n (%). JHS, Jackson Heart Study; MESA, Multi-Ethnic Study of Atherosclerosis; ARIC, Atherosclerosis Risk in Communities; WHI, Women’s Health Initiative; BMI, body mass index.

|  | Females | Males |
| --- | --- | --- |
| **Discovery** |  |  |
| JHS |  |  |
| n | 1042 | 620 |
| BMI: mean (sd) | 32.88 (7.30) | 30.17 (6.04) |
| age: mean (sd) | 56.22 (12.20) | 55.4 (12.36) |
| smoking status<br>(never/past/current) | 754 (72.4%)/ 170 (16.3%)/ 118 (11.3%) | 323 (52.1%)/ 176 (28.4%)/ 121 (19.5%) |
| MESA |  |  |
| n | 103 | 76 |
| BMI: mean (sd) | 31.73 (5.89) | 28.97 (4.94) |
| age: mean (sd) | 60.94 (9.50) | 60.67 (9.76) |
| smoking status<br>(never/past/current) | 59 (57.3%)/ 27 (26.2%)/ 17 (16.5%) | 26 (34.2%)/ 33 (43.4%)/ 17 (22.4%) |
| **Replication** |  |  |
| ARIC |  |  |
| n | 1458 | 859 |
| BMI: mean (sd) | 31.36 (6.66) | 28.05 (5.05) |
| age: mean (sd) | 56.73 (5.81) | 56.89 (6.03) |
| smoking status<br>(never/past/current) | 797 (54.7%)/ 354 (24.3%)/ 307 (21.0%) | 223 (26.0%)/ 333 (38.7%)/ 303 (35.3%) |
| WHI |  |  |
| n | 1129 | X |
| BMI: mean (sd) | 31.38 (6.39) | X |
| age: mean (sd) | 62.34 (6.74) | X |
| smoking status<br>(never/past/current) | 612 (54.2%)/ 379 (33.6%)/ 138 (12.2%) | X |

#### JHS (Jackson Heart Study)

JHS is a prospective population-based study of the high prevalence of common complex diseases among AAs in the Jackson, Mississippi metropolitan area. A total of 5306 self-identified AAs, aged between ages 20-95 years, were recruited during the baseline examination period in 2000-2004. A subset of these individuals (n=1756) had methylation measured from whole blood samples collected at baseline using the Illumina EPIC array (∼850K CpGs). BMI was calculated based on measured weight and standing height, and age, sex, and smoking status (past, current, never) were ascertained by self-report at baseline. The analysis sample consisted of n=620 males and n=1042 females with methylation data that passed quality control parameters and had complete BMI, age, sex, and smoking status information.

#### MESA (Multiethnic Study of Atherosclerosis)

MESA is a prospective population-based study of subclinical cardiovascular disease and its progression. A total of 6814 individuals, aged 45 to 84 years, were recruited from six U.S. communities (Baltimore City and County, MD; Chicago, IL; Forsyth County, NC; Los Angeles County, CA; New York, NY; and St. Paul, MN) between July 2000 and August 2002. Pre-specified recruitment plans identified four RE groups (White European American, African American, Hispanic American, and Chinese American) for enrollment, with targeted oversampling of minority groups to enhance statistical power. Methylation was measured on subset of individuals (n=1890) using whole blood samples collected at baseline (Illumina EPIC array). The subset of individuals with methylation data maintains the proportion of racial/ethnic groups observed in the entire cohort. BMI was calculated based on measured weight and standing height, and age, sex, and smoking status (past, current, never) were ascertained by self-report at baseline. For this analysis, only the subset of AA males (n=76) and females (n=103) with methylation data that passed quality control parameters and had complete BMI, age, sex, and smoking status information was used. Individuals missing any of this information was excluded.

#### ARIC (Atherosclerosis Risk in Communities Study)

The ARIC study is a multi-center prospective population-based study of atherosclerotic disease in bi-racial populations. Males and females were recruited from four communities: Forsyth County, NC; Jackson, MS; suburban areas of Minneapolis, MN; and Washington County, MD between 1987-1989. A total of 15,792 individuals, aged 45-64 years were examined at baseline. Individuals from Jackson that are also present in the JHS dataset were excluded from the analysis of the ARIC cohort. A total of 3407 individuals had methylation measured from whole blood samples collected at baseline using the Illumina Human Methylation 450k array. BMI was calculated from measured weight and standing height, and age, sex, and smoking status were ascertained by self-report at baseline. Only the subset of AA males (n=859) and females (n=1458) that had methylation data that passed quality control parameters, were not also present in the JHS dataset, and had complete BMI, age, sex, and smoking status information were included in this analysis.

#### WHI (Women’s Health Initiative)

WHI is one of the largest (n=161,808) studies of women’s health ever undertaken in the U.S. The women were recruited from 40 clinics across the U.S. between 1993-1998. Women enrolled range from ages 50-79 years, and 17% (∼26,000) identified with minority populations. A subset of individuals (n=2065) had methylation measured from whole blood collected at baseline using the Illumina Human Methylation 450k array. BMI, age, and smoking status (past, current, never) were ascertained by self-report at baseline. Only the subset of AA women (n=1189) with methylation data that passed quality control parameters and that had complete BMI, age, and smoking status information were included in this analysis.

### Exclusion criteria

#### Subjects

Individuals were excluded if they did not have methylation data or were missing >1% of methylation data or there was a mismatch between self-reported and biological sex.

#### Methylation

Methylation sites were excluded from analysis if they had <99% call rate, have previously been reported as cross-reactive, have a variance ±4 standard deviations from the mean variance, or are within 5bp of a single-nucleotide polymorphisms (SNPs) with a minor allele frequency (MAF) >1% for AAs (Zhou et al., 2017). Documentation of cross-reactive sites and sites near common SNPs for AAs can be found at https://zwdzwd.github.io/InfiniumAnnotation. After exclusion, there were 573,388 sites shared between JHS and MESA that were used for the discovery analysis set (individual CpG and DMR analyses). ARIC and WHI methylation were measured using the 450K Human Methylation array, and after exclusions there were 305,908 sites remaining that overlapped with sites present in the discovery set for replication testing of individual CpG associations.

#### Epigenome-Wide Association analyses

EWAS analyses were stratified by study and sex. For each study-sex stratum, BMI was used as the main predictor in the linear regression model. Methylation M-values were used as the outcome in the model. Prior to analysis, the M-values were adjusted for technical covariates (batch, plate, well, etc.) using the ComBat function in the sva R package v3.48.0 (Johnson et al., 2007). Covariates for each model included age, centered-age-squared ((*age –* mean(age))^2^), smoking status (never, past, current), the first ten genetic principal components, and estimated blood cell proportions using the Houseman method (Houseman et al., 2012). If a study had participants from multiple sites, then study site was also included as a covariate in the model.

The general linear regression model used was:

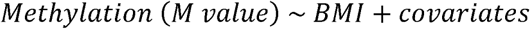

After running the stratified linear regression analyses, the results were adjusted for bias and inflation using the BACON R package v1.28.0 (van Iterson et al., 2017). BACON is a Bayesian method developed for transcriptomic and epigenomic association studies, and it has been shown to perform better in these scenarios than methods devised for genomic association studies such as genomic control. The Bonferroni-corrected significance threshold was p=⍰18.72 ×⍰110^–8^ to account for multiple testing (0.05/573,388 tests). This threshold is more stringent than the commonly used Benjamini-Hochberg FDR-adjustment and is comparable to the recommended significance threshold for EWAS using the EPIC array (Mansell et al., 2019).

Summary statistics from the study- and sex-stratified linear regression analyses were meta-analyzed using the inverse-variance weighted fixed-effects method implemented in the command line tool METAL (version “2020-05-05”) (Willer et al., 2010). JHS and MESA AA male and female results were meta-analyzed for discovery of BMI-associated CpGs. WHI and ARIC AA male and female results were meta-analyzed for replication of results identified in the JHS and MESA analysis. Of the 151 novel epigenome-wide significant CpGs from the discovery set, only 29 were present in the ARIC-WHI 450k array data for replication testing. Replication was defined as sites having the same direction of the beta coefficient in the discovery and replication sets and a Bonferroni adjusted significance threshold of p= 1.72 x 10^-2^ (0.05/29 tests).

### Replication of previously reported CpGs

Previously reported CpGs associated with BMI were downloaded from the EWAS Atlas Open Platform database. In total, there were 2435 unique CpG sites associated with BMI from 31 studies that were conducted primarily in cohorts of European genetic ancestry. Four of the 31 studies included the AA subset from ARIC. At the time of this study there were only eight published EWAS for obesity-related traits conducted in AA-only samples, five of which also included the AA subset from ARIC (Akinyemiju et al., 2018; Demerath et al., 2015; Dhana et al., 2018; Do et al., 2023a; Mendelson et al., 2017; Sun et al., 2019; Taylor et al., 2023; Wang et al., 2018). The studies by Akinyemiju et al., Do et al., Sun et al., and Wang et al. were not included in the EWAS Atlas database at the time of this analysis. From these eight studies, there were a total of 278 unique CpG sites identified or replicated in AA, 124 of which were not present in the EWAS Atlas BMI data. We considered a significant (p<8.72x10^-8^) CpG in our discovery study to be replicating a previously reported CpG result if the direction of effect in our study was consistent with at least one previously reported study with a significant finding for that CpG.

### Enrichment Analyses

All enrichment analysis were performed using the EWAS Open Platform ATLAS Toolkit (https://ngdc.cncb.ac.cn/ewas/toolkit). GO and KEGG enrichment is based on the ‘gometh’ function from the missMethyl R package, which accounts for differing numbers of probes per gene present based on the methylation array type (450k or 850k) and for CpGs that have annotations with multiple genes (Phipson et al., 2015). Trait enrichment is performed using a weighted Fisher’s exact test to calculate the co-occurrence probability of queried CpGs with CpGs that have previously been reported as significant with a trait from studies submitted to the EWAS Atlas database.

### DMR (Differentially Methylated Regions) Analysis

DMR analysis was performed using the command-line tool Comb-p with the summary statistics for all 573,388 CpGs tested from the discovery EWAS (Pedersen et al., 2012). The Comb-p method uses a sliding-window approach that accounts for spatial correlation and uneven distribution of sites across the genome to identify and assign significance to regions with enrichment. A p-value of <8.7x10^-8^ was required to start a region, and a region would be extended if another significant site was found within 1000bp of the previous site.

### Mediation Analyses

Previously reported BMI-associated SNPs were identified using the summary statistics from BMI GWAS conducted in the Genetic Investigation of ANthropometric Traits (GIANT) (https://portals.broadinstitute.org/collaboration/giant/images/e/e2/Meta-analysis_Locke_et_al%2BUKBiobank_2018_top_941_from_COJO_analysis_UPDATED.txt.gz). Cis-SNPs were defined as SNPs within 1Mbp of BMI-CpGs identified in the discovery EWAS. We identified 136 cis-BMI-SNPs using the Genomic Ranges R package v1.53.1 (Lawrence et al., 2013). Then, we performed association testing of the cis-BMI-SNPs and BMI in the discovery cohorts (JHS and MESA). Since no statistically significant associations were observed between the cis-BMI-SNPs and BMI in our discovery sample at the Bonferroni adjusted p-value threshold p=3.68 x 10^-4^ (0.05/136 tests), no mediation testing was performed.

Sensitivity Analyses:

Sensitivity analyses were performed to assess if socioeconomic factors, which could have significant effects on methylation and obesity, impacted our results. To asses this, we conducted sex-stratified association testing of the top 208 methylation sites with BMI using the same linear regression models and meta-analysis methods as outlined above, but we additionally included educational attainment level (less than high school/GED/post-high school) and family income (<5k, 5-7k, 8-12k, 12-16k, 16-20k, 20-25k, 25-35k, 35-50k, 50-75k, 75-100k, >100k) as covariates. There were 220 individuals (156 females, 64 males) from JHS who did not have complete educational attainment or family income data and were excluded from the sensitivity analysis. All MESA individuals who were included in the discovery analysis had complete educational attainment and family income information.

### Software

Analyses were performed using the R programming language v4.1.3, the EWAS Atlas Toolkit (https://ngdc.cncb.ac.cn/ewas/toolkit), and the command-line tools METAL and Comb-p (Austin, 1990; Pedersen et al., 2012; Willer et al., 2010). The code for performing epigenome-wide association testing can be found at https://github.com/k2ferrier/Methylation-Data-Processing.

## 3. RESULTS

### Discovery

After meta-analysis of sex-stratified EWAS of BMI in JHS and MESA AAs, 208 methylation-sites surpassed epigenome-wide significance (p< 8.72 x 10^-8^, Figure 2, Supplementary Table 1) in our discovery set. These 208 sites annotated to 216 genes; 23 CpGs had 2+ gene annotations and 29 CpGs had no gene annotations. Results showed that 151 of the 208 associations were novel BMI-associated CpGs annotated to 154 genes (24 CpGs with 2+ gene annotations and 27 CpGs with no gene annotations). The top novel CpG site was cg02370334 (p=1.10 x 10^-21^), located downstream of *JAK3* (Janus Kinase 3). One other CpG in *JAK3*, cg20959703, was also significant (p=9.90x10^-15^) in our results. Interestingly, this gene has not been associated with obesity in previous EWAS or GWAS, but differential expression of *JAK3* was reported in a TWAS of BMI in the White and Hispanic American subsets from MESA and showed suggestive significance in the subset of AAs (Vargas et al., 2023). Other CpGs in or near 34 of the 154 annotated genes have previously been reported in the BMI EWAS in the EWAS Atlas. Fifteen of the novel sites were in or near genes that have previously been associated with BMI in GWAS, including: *ADARB1*, *ARAP1*, *CYP7B1*, *DLEU1*, *EGLN3*, *IGF1R*, *MAD1L1*, *MARK4*, *MYO1E*, *NRXN2*, *RAPGEF4*, *RNF216*, *SRR*, and *TNIK*.

**Figure 2:**
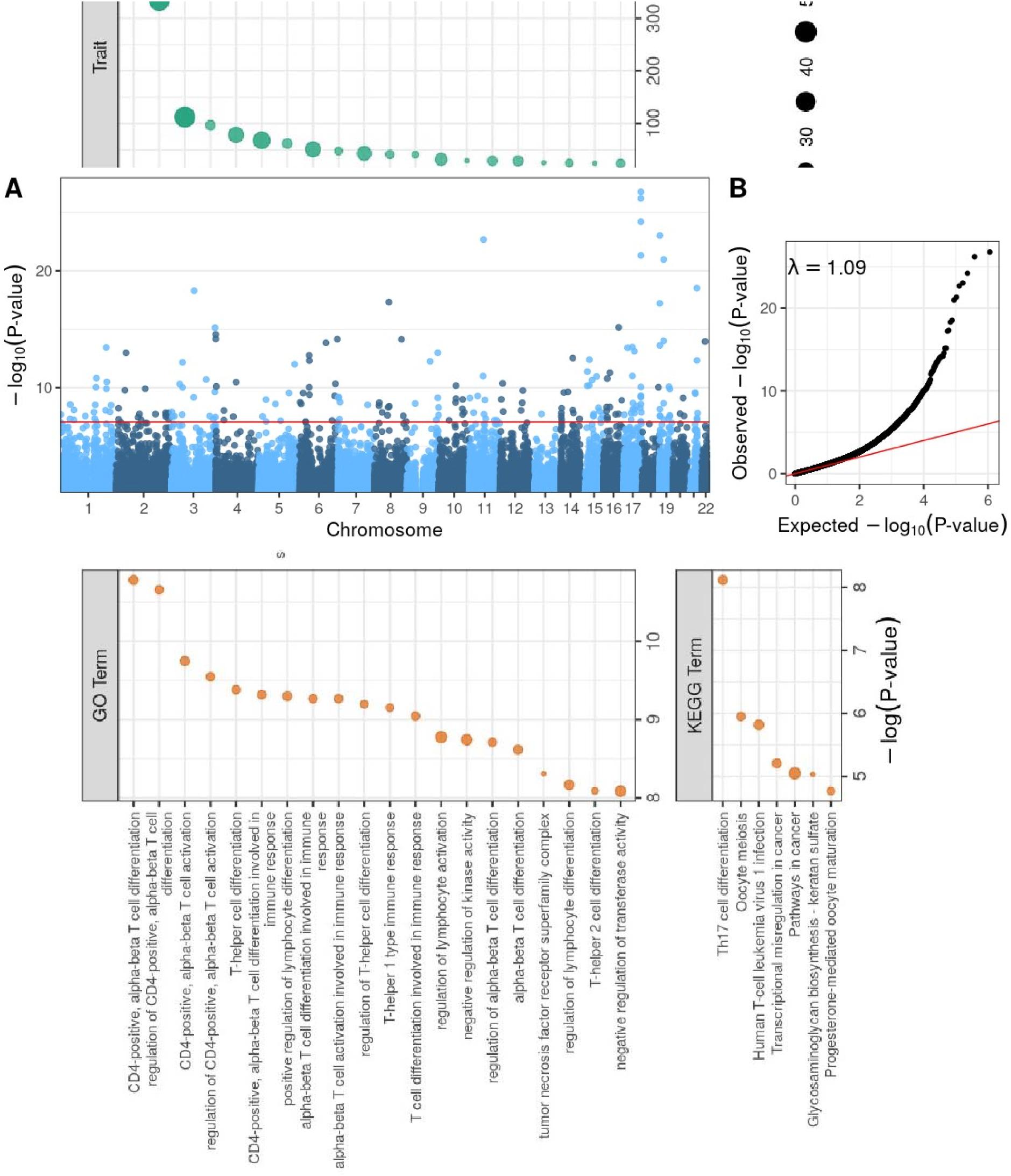
Discovery BMI EWAS meta-analysis results in African Americans. A) Manhattan plot of the association between DNA methylation and BMI in African Americans. B) QQ-plot of expected vs observed results. The genomic inflation factor after BACON-adjustment (A)= 1.09.

The two cohorts used for replication analysis (ARIC and WHI) had methylation data measured on the smaller 450k Human Methylation array, so only 29 of the 151 novel sites discovered could be tested for replication. All 29 CpGs had beta-coefficients in the same direction in the replication results as the discovery results, but only 20 showed statistical significance (p<1.72 x 10^-2,^ Supplementary Table 2). The 20 replicated novel sites annotated to 25 genes; 7 CpGs had 2+ gene annotations, 3 had no gene annotations (Supplementary Table 3). The top replicated novel site was cg12269535 located near *SRF* (Serum Response Factor). *SRF* has not been associated with any obesity-related EWAS in the EWAS Atlas, but SNPs in a paralog of *SRF* (*MEF2D*) have been reported in BMI GWAS in the GWAS Catalog. Other CpGs in or near 5 of the 25 genes have been reported in BMI EWAS in the EWAS Atlas (e.g., *PHACTR2, CALHM1, IGF1R, LAP,* and *ADARB1*), including cg26712428 in the south shore of *CALHM1* (Calcium Homeostasis Modulator 1) which was also significant in our results (p=1.38 x 10^-17^). *CALHM1* is a gene for a calcium ion channel that processes amyloid-beta precursor proteins and has been associated with Alzheimer’s disease (Dreses-Werringloer et al., 2008). The potential role of this gene in obesity is unclear, but genetic loci in this gene have been associated with body height in previous GWAS (Okbay et al., 2022; Yengo et al., 2022). Four of the replicated novel CpGs were in or near three genes which have previously been associated with BMI in GWAS studies (*ADARB1*: cg22635096, *IGF1R*: cg23900712, *MYO1E*: cg24263283, cg08423142).

### Replication of previously reported CpGs

Of the 208 sites significantly associated with BMI, 45 have previously been reported to be associated with BMI in the EWAS Atlas (https://ngdc.cncb.ac.cn/ewas/). Effect size summary statistics were only available for 42 of the 45 previously reported sites, but our results showed beta-coefficients with the same direction as at least one of the previously reported results for all 42 sites (100%). Four sites had multiple summary statistics reported that included both negative and positive beta coefficients (cg04816311, cg08857797, cg24531955, and cg18181703) in the EWAS Atlas. Twelve additional CpGs were previously reported in studies of only AAs and not included in the EWAS Atlas. Our results showed that all twelve sites replicated the previously reported AA summary statistics.

Of note, 4 CpGs near *SOCS3* (Suppressor of cytokine signaling 3) identified by Taylor et al. in an AA BMI EWAS were replicated in this study: cg11047325 (p=1.74x10^-27^), cg13343932 (p=4.83 x 10^-22^), cg10508317 (p=8.79 x 10^-11^), and cg18181703 (p=6.37 x 10^-27^). SOCS3 is a negative regulator of cytokine signaling and has been associated with Atopic Dermatitis 4 and overnutrition (Yin et al., 2015). Differential methylation of cg18181703 and cg10508317 has been observed in European, AA, and South Asian populations in BMI EWAS in the EWAS Atlas, but associations of cg11047325 and cg13343932 have only been identified in cohorts of AAs so far (Taylor et al., 2023). In addition to the 2 *SOCS3* CpGs, there were 9 other CpGs identified in our analysis that have only been reported in BMI EWAS of AAs: cg19748455 (*LINC01993*), cg19758958 (*AHNAK*), cg24382141 (*PSKH1*), cg03770138 (*AL162417.1*;*RALGDS*), cg27540367 (*TGFB1*), cg04610187 (*AC061992.2*), cg11793449 (*AC087645.2*), cg20803896, cg04986899 (*XYLT1*) (Do et al., 2023b; Sun et al., 2021; Taylor et al., 2023) *Differentially Methylated Region (DMR) Analyses:*

DMR analyses of the 208 significant sites from the discovery set showed 54 significant regions that contained two or more significant individual sites (Supplementary Table 3). Several of these regions are proximal to, or overlap, genes that have previously been reported in association with anthropometry-related traits in genetics or epigenetics studies, including *SOCS3*, *SBNO2*, *ABCG1*, *WDR8*, *B3GALT4, TGFB1, CYP7B1*, *ZAP70*, *C21ORF62*, and *MAD1L1.* Two DMRs were proximal to the enhancer region of gene *JAK3*, which showed significant differential expression in a previously published multi-ethnic BMI meta-analysis of MESA individuals, but was not significant in the AA subset alone (Vargas et al., 2023). The first and third most significant DMRs from this analysis include five CpGs in the proximal enhancer region (z Sidak p= 8.07 x 10^-61^) and three CpGs in the promoter region (z p=9.86 x 10^-30^) of *SOCS3*. There were two significant DMRs from our results in the distal (z Sidak p=2.20 x 10^-26^)and proximal (z Sidak p=6.06 X 10^-9^) enhancer regions of *ABCG1* (ATP binding cassette subfamily G member 1), which is a gene that has shown significant associations between differential methylation, gene expression, and BMI in a previous study (Mendelson et al., 2017).

### Trait, Gene, and Pathway and Enrichment Analyses

Gene Ontology (GO) enrichment of the 208 CpGs from the discovery set showed statistically significant enrichment with activation or differentiation of immune system related cell type pathways (Figure 3A). The strongest GO enrichment associations were as follows: with the CD-4 positive, alpha-beta T cell differentiation pathway (p=2.07 x 10^-5^); regulation of CD4-positive, alpha-beta T cell differentiation (p=2.35 x 10^-5^); CD-4 positive, alpha-beta T cell activation (p=5.84 x 10^-5^); and regulation of CD4-positive, alpha-beta T cell activation (p=7.14 x 10^-5^). Other GO enrichment analysis results are listed in Supplementary Table 4. KEGG pathway enrichment analysis showed statistically significant enrichment with Th17 cell differentiation (p=2.98 x 10^-4^), oocyte meiosis (2.60 x 10^-3^), Human T-cell leukemia virus 1 infection (p=2.97 x 10^-3^), transcriptional misregulation in cancer (p=5.46 x 10^-3^), pathways in cancer (p=6.39 x 10^-3^), glycosaminoglycan biosynthesis—keratan sulfate (p=6.52 x 10^-3^), and progesterone-mediated oocyte maturation (p=8.48 x 10^-3^) (Supplementary Table 5).

**Figure 3:** Pathway and Enrichment Analyses of significant BMI-CpGs in African Americans. A) Trait enrichment analysis. B) KEGG Pathway enrichment analysis. C) GO enrichment analysis.

The top five traits associated with the epigenome-wide significant CpGs from the discovery set in a trait enrichment analysis were: Crohn’s disease (n=58 CpGs, p=2.49 x 10^-185^, OR=142.57), BMI (n=44 CpGs, p= 1.98 x 10^-145^, OR=147.99), aging (n=47, p=1.71 x 10^-49^, OR=7.69), inflammatory bowel disease (IBS) (n=7 CpGs, p=8.39 x 10^-43^, OR=5443.19), and obesity (n=23 CpGs, p=7.67 x 10^-35^, OR=14.63) (Figure 3C). The odds ratio for IBS is comparatively much higher than the other traits due to there being many fewer IBS-CpGs present in the database (n=14) versus the other traits (n>1700). A table of all significantly associated traits can be found in Supplementary Table 6.

### Sex-Specific Results

In the subset of AA females from JHS and MESA there were 88 epigenome-wide significant results. Our results showed that 77 of these sites were also significant in the meta-analysis of results from both males and females in the same cohorts. The subset of AA males from JHS and MESA had seven epigenome-wide significant results, four of which were also significant in the sex-combined results. Only one site (cg13274938) was significant in both the female- and male-specific results. Overall, we observed more sites significant in the female stratum than the male stratum (Figure 4A). While these results could be due to true biological differences in sex, the differences in sample size between the male and female subsets could explain why fewer significant sites were detected in the male subset. Additionally, the estimated effect sizes (beta coefficient from the linear regression summary statistics) were slightly more extreme for sites significant in the female-stratum than the male stratum (Figure 4B). Future sex stratified EWAS of BMI with a balanced female to male ratio will be necessary to validate these findings. All sex-specific results can be found in Supplementary Tables 7 and 8.

**Figure 4:**
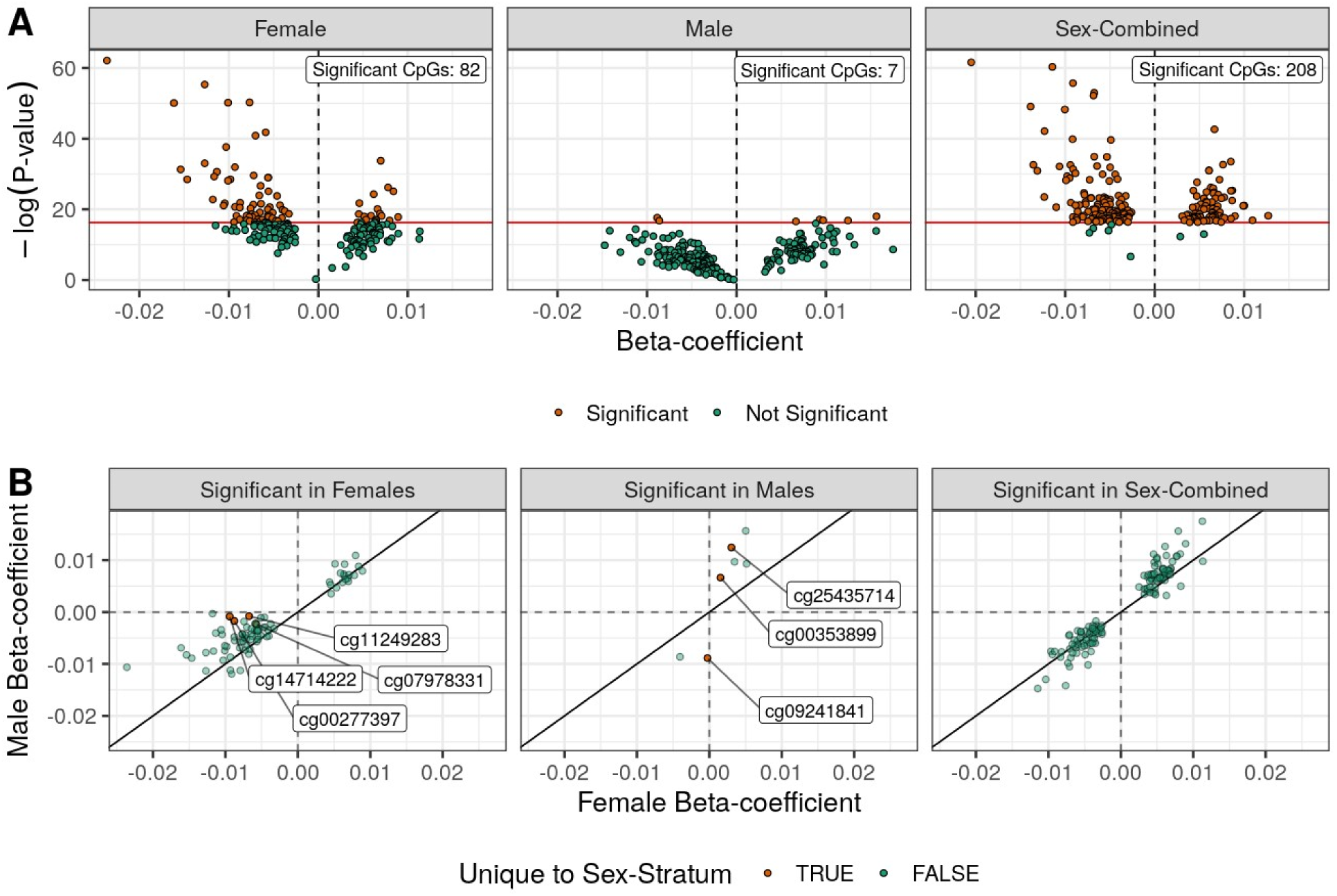
Comparison of sex-specific versus meta-analyzed EWAS results. (A) Volcano plots of the beta-coefficients and p-values for the epigenome-wide significant sites across the sex-specific and sex stratified meta-analysis results (n=215). The leftmost plot shows the results for these sites in the female stratum, the middle plot shows the results for the male stratum, and the rightmost plot shows the results for the sex stratified meta-analysis (sex-combined). (B): Scatter plots of the male versus the female beta-coefficients for the epigenome-wide significant sites from each sex stratum and the sex stratified meta-analysis (sex-combined). The leftmost plot shows the results for the 82 sites significant in the female stratum with labels for the four sites which were unique to the female stratum analysis. The middle plot shows the results for the 7 sites significant in the male stratum with labels for the three sites that were unique to the male stratum analysis. The rightmost plot shows the results for the 208 significant sites from the sex stratified meta-analysis.

### Sensitivity Analyses

Inclusion of educational attainment level and family income did not significantly impact the direction or magnitude of the beta-coefficients or p-values for any of the 208 significant sites from the discovery analysis.

## 4. Discussion

In this study, we performed a sex-stratified epigenome-wide association meta-analysis of BMI using methylation data measured by the Illumina EPIC 850k array on PBMCs or whole blood samples collected from 1,841 AA participants of JHS and MESA. From our discovery analysis, we identified 208 methylation sites (CpGs) with differential methylation significantly associated with BMI in AAs. We found that 151 of the 208 significant BMI-CpGs were novel in their association with BMI. We identified 57 associated CpGs that were previously reported in other BMI EWAS (mostly from 450k array data); 45 from BMI EWAS in the EWAS Atlas and an additional 12 from AA obesity-related EWAS not included in the EWAS Atlas database at the time of this study. A majority of the novel CpGs identified by our analysis were not present on the older 450k methylation array (n=129), which was the most commonly used platform for methylation interrogation until 2016 when the EPIC 850k panel was released. In an independent dataset of 3,506 AAs from ARIC and WHI with 450k methylation array data, we were able to assess replication for 29 novel sites (65.5%) that overlapped between the 850K and 450K array data. The replication results had concordant direction of effects for all 29 sites compared with the discovery results, but only 20 were statistically significant (p<1.72x10^-2^). Of the genes that annotated to the 20 replicated novel CpGs: 3 genes (*PHACTR2, CALHM1,* and *LAP*) had other nearby CpGs that have previously been reported in BMI EWAS in the EWAS Atlas; 1 gene (*MYO1E*) had SNPs associated with BMI in the GWAS Atlas; and 2 genes (*IGF1R* and *ADARB1*) have previously been implicated with BMI in both EWAS and GWAS.

Since methylation status can be influenced by both genetics and the environment, it is possible there are environmental factors that are driving the differential methylation. We performed sensitivity analysis on all significant CpGs that controlled for educational attainment and total household income as proxies for socioeconomic status, and the results were consistent in terms of the effect size and significance. Furthermore, there were no SNPs in cis-with any of the top BMI-CpGs from our results that showed a significant genetic association with BMI in our discovery cohorts, so no mediation testing could be performed to investigate the relationship between the genetics, methylation, and BMI. It is plausible that reverse causality is driving these BMI-CpG associations (i.e., BMI is directly influencing methylation levels rather than methylation levels directly influencing BMI). Future investigations between the genome, epigenome, and environmental factors will be necessary to validate and disentangle the causal relationship between these novel BMI-CpGs in the development of obesity in AAs.

DMR analysis of the top sites from the discovery analysis showed 54 significantly differentiated regions in association with BMI. These regions were proximal or included genes that have previously been implicated with anthropometry-related traits and obesity-relevant processes, including *SOCS3*, *SBNO2*, *ABCG1*, *WDR8*, *B3GALT4, TGFB1, CYP7B1*, *ZAP70*, *C21ORF62*, and *MAD1L1*. Pathway and gene enrichment analysis showed enrichment for genes and pathways involved in immune system cell differentiation and activation. This is consistent with previous studies of genetic and epigenetic markers of obesity in AA (Demerath et al., 2015; Dhana et al., 2018; Do et al., 2023a; Sun et al., 2019; Wang et al., 2018). Trait enrichment analysis revealed strong associations with autoimmune and inflammatory diseases, aging, and obesity.

Our results show several top CpGs (including some novel) and DMRs that are proximal to or include *SOCS3*, which is one of the genes targeted in the IL-6/JAK/STAT pathway*. SOCS3* also showed differential expression in association with BMI in the AA subset of the MESA cohort (Vargas et al., 2023). Furthermore, a previous EWAS of obesity showed a significant interaction between methylation of cg18181703 in *SOCS3* and cumulative stress on BMI in a cohort of healthy EA and AA participants (Xu et al., 2018). Several of our other top CpGs and DMRs have gene annotations that are related to the IL-6 (Interleukin 6)/JAK (Janus kinase)/STAT (signal transducer and activator of transcription) pro-inflammation pathway. IL-6 is a cytokine that is produced in response to infections, tissue injury, and can be activated by acute stress (Qing et al., 2020). Continuous production of IL-6 causes a transition from acute to chronic inflammation with immune responses that proliferate inflammation. The JAK/STAT pathway is one of the signaling mechanisms activated by IL-6 and leads to expression of STAT-dependent IL-6 target genes. Another target gene of the IL-6/JAK/STAT pathway that was identified by our analysis is *JAK3*. While this gene is novel in EWAS of anthropometry-related traits in humans, loss of *JAK3* has been shown to cause increased body weight and chronic low-grade inflammation in mice (Kumar et al., 2022). Differential expression of *JAK3* was significant in the BMI meta-analysis across race/ethnic groups in MESA. Since methylation status can only be used to infer gene expression, further testing of this gene in AAs will be necessary to validate if differential methylation is causing changes in *JAK3* expression in this population. Other IL-6/JAK/STAT target genes or genes that interact with this pathway that showed significant differentially methylated CpGs and/or DMRs in our results include *BCL3*, *BCL6*, *SBNO2*, and *TGFB1*.

The results of these analyses support that there are potentially obesity associated epigenetic markers with increased signals in AAs. However, we note that most of our novel findings are on the non-overlapping set of CpGs not included on the older 450k array and need to be confirmed in independent studies of AA with 850k array data. Future investigations will also need to be conducted to assess whether these CpGs are truly involved in obesity pathogenesis and/or if these associations are being driven by unaccounted for genetic or environmental factors. Additionally, BMI is an indirect measurement of fat mass that is non-invasive and easy to obtain but does not accurately describe body fat composition. Studies of directly measured adiposity could improve power to discern sex- and/or RE-specific effect-size estimates or markers of obesity. It will also be important to validate these results in studies with methylation measurements taken from adipose tissue versus whole or peripheral blood, given the known tissue specificity of methylation patterns. Another notable limitation of this study is that analyses were done using cross-sectional data. Since methylation patterns are influenced by environmental factors and can change over time, studies incorporating longitudinal data will be necessary to clarify temporally stable and causal relationships between genetics, epigenetics, environmental factors, and the development of obesity.

To summarize, in this study we performed sex-stratified EWAS meta-analyses of BMI in AAs and identified 208 epigenome-wide significant CpGs and 54 statistically significant DMRs. We observed high replication of results for CpGs previously reported in multi-ethnic and AA-specific BMI EWAS and were able to replicate 20 of the 151 novel CpGs in a separate sample of AAs. Trait, gene, and pathway enrichment analyses support that chronic inflammation may be a key contributing factor in the etiology of obesity in AAs. Future research targeting these genes or pathways in other samples of African Americans could aid in the development of personalized medicine approaches for a demographic of people disproportionately burdened by obesity and underrepresented in medical research.

## Data Availability

All data produced in the present study are available upon reasonable request to the authors.

## ACKNOWLEDGEMENTS

Molecular data for the Trans-Omics in Precision Medicine (TOPMed) program was supported by the National Heart, Lung and Blood Institute (NHLBI). Methylomics for “NHLBI TOPMed: Multi-Ethnic Study of Atherosclerosis (MESA)” (phs001416.v3.p1) was performed at Keck MGC (HHSN268201600034I). Genome sequencing was performed at the Broad Institute of MIT and Harvard (3U54HG003067-13S1). Centralized read mapping and genotype calling, along with variant quality metrics and filtering were provided by the TOPMed Informatics Research Center (3R01HL-117626-02S1 and HHSN268201800002I). Phenotype harmonization, data management, sample-identity QC, and general study coordination, were provided by the TOPMed Data Coordinating Center (3R01HL-120393-02S1), and TOPMed MESA Multi-Omics (HHSN2682015000031/ HHSN26800004). MESA and the MESA SHARe project are conducted and supported by the National Heart, Lung, and Blood Institute (NHLBI) in collaboration with MESA investigators. Support for MESA is provided by contracts HHSN268201500003I, N01-HC-95159, N01-HC-95160, N01-HC-95161, N01-HC-95162, N01-HC-95163, N01-HC95164, N01-HC-95165, N01-HC-95166, N01-HC-95167, N01-HC-95168, N01-HC-95169, UL1-TR-001079, UL1-TR000040, UL1-TR-001420, UL1-TR-001881, and DK063491. The MESA Epigenomics & Transcriptomics Study was funded by NIA grant 1R01HL101250-01 to Wake Forest University Health Sciences. The authors thank the other investigators, the staff, and the participants of the MESA study for their valuable contributions. A full list of participating MESA investigators and institutes can be found at http://www.mesa-nhlbi.org.

The Jackson Heart Study (JHS) is supported and conducted in collaboration with Jackson State University (HHSN268201800013I), Tougaloo College (HHSN268201800014I), the Mississippi State Department of Health (HHSN268201800015I) and the University of Mississippi Medical Center (HHSN268201800010I, HHSN268201800011I and HHSN268201800012I) contracts from the National Heart, Lung, and Blood Institute (NHLBI) and the National Institute on Minority Health and Health Disparities (NIMHD). The authors also wish to thank the staff and participants of the JHS. The views expressed in this manuscript are those of the authors and do not necessarily represent the views of the National Heart, Lung, and Blood Institute; the National Institutes of Health; or the U.S. Department of Health and Human Services.

The PAGE Study is funded by the National Human Genome Research Institute with co-funding from the National Institute on Minority Health and Health Disparities. Assistance with data management, data integration, data dissemination, genotype imputation, ancestry deconvolution, population genetics, analysis pipelines and general study coordination was provided by the PAGE Coordinating Center (NI-HU01HG007419). PAGE was also funded by grants R56HG010297 and R01HG010297. PAGE data and materials included in this report were funded through the following studies and organizations: The WHI program is funded by the National Heart, Lung, and Blood Institute, National Institutes of Health, and US Department of Health and Human Services through contracts 75N92021D00001, 75N92021D00002, 75N92021D00003, 75N92021D00004 and 75N92021D00005. The authors thank the WHI investigators and staff for their dedication and the study participants for making the program possible. A listing of WHI investigators can be found at https://www.whi.org/researchers/Documents%20%20Write%20a%20Paper/WHI%20Investigator%20Long%20List.pdf. Data from the WHI are available on request at https://www.whi.org/researchers/SitePages/Write%20a%20Paper.aspx. The Atherosclerosis Risk in Communities (ARIC) study has been funded in whole or in part with Federal funds from the National Heart, Lung, and Blood Institute, National Institutes of Health, Department of Health and Human Services (contract numbers HHSN268201700001I, HHSN268201700002I, HHSN268201700003I, HHSN268201700004I and HHSN268201700005I). ARIC DNAm funding was also provided by the American Recovery and Reinvestment Act of 2009 5RC2HL102419 and by R01-NS087541The authors thank the staff and participants of the ARIC study for their important contributions. Data from the ARIC study are available on request at https://www2.cscc.unc.edu/aric/distribution-agreements.

